# Intact finger representation within primary sensorimotor cortex of Musician’s Dystonia

**DOI:** 10.1101/2021.12.01.21266860

**Authors:** A Sadnicka, T Wiestler, K Butler, E Altenmüller, MJ Edwards, N Ejaz, J Diedrichsen

## Abstract

Musician’s dystonia presents with a persistent deterioration of motor control during musical performance. A predominant hypothesis has been that this is underpinned by maladaptive neural changes to the somatotopic organisation of finger representations within primary somatosensory cortex. Here, we tested this hypothesis by investigating the finger-specific activity patterns in the primary somatosensory and motor cortex using functional MRI in nine musicians with dystonia and nine healthy musicians. A purpose-built keyboard device allowed functional MRI characterisation of activity patterns elicited during passive extension and active finger presses of individual fingers. We analysed the data using both traditional spatial analysis and state-of-the art multivariate analyses. Our analysis reveals that digit representations in musicians were poorly captured by spatial measures. An optimised spatial metric found clear somatotopy but no difference in the spatial geometry between fingers. Representational similarity analysis was confirmed as a highly reliable technique and more consistent than all spatial metrics evaluated. Significantly, the dissimilarity architecture was equivalent for musicians with and without dystonia and no expansion or spatial shift of digit representation maps were found in the symptomatic group. Our results therefore suggest that the neural representation of generic finger maps in primary sensorimotor cortex is intact in Musician’s dystonia. These results are against the idea that task-specific dystonia is associated with a distorted hand somatotopy and suggests that task-specific dystonia is due to a higher order disruption of skill encoding. Such a formulation can better explain the task-specific deficit and offers mechanistic insight for therapeutic interventions.

## Introduction

Task-specific dystonia is a form of isolated dystonia that presents with a selective motor impairment during the performance of a specific skill.^1^ For example, in Musician’s dystonia there is normal use of the fingers for writing or typing yet the individual is unable to fluently co-ordinate the same fingers during musical performance.^2^ The highest relative prevalence of task-specific dystonia is seen within professional musicians with 1-2% affected.^3^ For many this is the end of performance careers, with a devastating impact both the individual and their contribution to our cultural society.^1, 4, 5^

A highly influential animal model of task-specific dystonia has dominated research over the last two decades.^6^ In this model, two monkeys were trained to perform rapid, repetitive, highly stereotypic grasping movements until a painful forearm syndrome developed and motor performance deteriorated significantly.^6^ Subsequent electrophysiological mapping of the representations of the hand within primary somatosensory cortex revealed a 10-20 fold increase in the size of the sensory receptive fields of neurons. In addition, there was the breakdown of normally segregated areas, for example, the representation of the dorsal and palmar aspects of hand were found to be overlapping. Motivated by this primate study, electrophysiological and imaging studies in humans have also provided evidence of distorted finger representations primary somatosensory cortex (S1) in task-specific dystonia.^7-14^ Therefore, one hypothesis is that the pathophysiology of task-specific dystonia is caused by a distorted somatotopy of the hand in S1.

However, alternative disease models have also been proposed as altered representations within primary cortical regions are unable to explain many clinical features. For example, such a model cannot explain task-specificity. Abnormal digit representations in S1 would predict a general deficit of *any* task in the implicated fingers as the representation or encoding of sensory information at its lowest level is corrupted. Additionally, even within the affected task, dystonia is varied in its manifestations. Some musicians only experience deficits only within a particular sequence with precise spatial and temporal features (such as playing tremolo on the guitar, or an ascending scale rather than a descending scale on the piano). Such features represent complex and more abstract features of movement that are encoded within higher order control regions of the motor hierarchy such as the premotor and parietal cortices.^15^

Recent advances in the analysis of distributed brain activity patterns have led to a novel view of how the hand and fingers are represented in sensorimotor cortex.^16^ Traditional approaches have stressed the orderly spatial mapping of body parts to different regions of the brain, reinforcing the notion of the iconic homunculus. In contrast to this notion of a discrete, orderly layout, both for hand and the entire body, functional MRI (fMRI) studies in healthy individuals have shown substantial overlap between cortical areas activated by different body parts, often showing complex layouts with multiple peaks of activation both in M1 and S1.^17, 18^ This does not equate to a simple segregated ordering of finger-specific activity as suggested by historical depictions of the homunculus. Furthermore, in health, such patterns are highly variable across individuals.^17^ Novel analysis methods such a representational similarity analysis have revealed that, although the actual spatial layout is variable across individuals, the relative overlap, or similarity, is highly preserved. This invariant organisation, with the thumb having the most unique representation and most distinct from the ring finger, can be explained by the statistics of finger movements in everyday activities.^17^ Thus, it appears that representations in sensorimotor cortex, rather than being dictated by a fixed spatial layout, arise from a mapping process of every-day actions onto the surface of the neocortex.^19^

We were therefore motivated to re-explore task-specific dystonia using updated neuroimaging methods to test the hypothesis of an altered finger map in S1 as the neural core correlate of the disease. To address this fundamental question, we built a robotic device that allowed fMRI characterisation of activity patterns elicited during a passive condition in which individual fingers were passively lifted by pneumatic pistons within the keys of the robot and an active condition in which individual fingers pressed down on piano like keys. Our analysis first compared the reliability of more traditional, spatial metrics of finger representations to the newer pattern-similarity based measures (Summary of Experiments, Table 1). We then compared musicians with task-specific dystonia to a healthy control group. The hypothesis of an altered map of finger representations would certainly need to predict an altered similarity structure of the underlying patterns. Specifically, the idea of increasing overlap and fusion of finger representations predicts a decreasing dissimilarity of the patterns of the affected fingers in Musician’s dystonia. Alternatively, a preserved organisation of basic finger representations in S1 and M1 would suggest that task-specific dystonia is due to alterations in other areas.

**Table 1.** Summary of Analyses. Each question is followed by the analytical approach used to address the query, the key result and the corresponding figure.

| Question | Approach | Result | Figure |
| --- | --- | --- | --- |
| How well do spatial fMRI metrics characterise the finger representation in S1? | Split-half reliabilities for a range of different metric were compared | Digit representations are poorly captured by single spatial measures | 2, 3 |
| Does the spatial geometry differ between healthy musicians and musicians with dystonia? | Euclidean distance between fingers using highest performing softmax COG were calculated for each individual. | Both groups showed clear somatotopy yet the spatial geometry was indistinguishable | 4 |
| Does multivariate pattern analysis better quantify finger representations in S1? | Representation similarity analysis (RSA) assessed the similarity between activity patterns for different fingers. | Split-half reliabilities for RSA were highly reliable in S1 (>0.8) and more consistent than all spatial metrics evaluated. | 3 |
| Is the representation structure in S1 altered in musicians' dystonia? | Dissimilarity between each finger pair was computed using the cross-validated Mahalanobis distance. | Dissimilarity architecture equivalent for patient and controls in S1 | 5 |
| Are there overall differences in the location or spatial extent of digit representations? | Statistical comparison of surface-based searchlight maps | No significant expansion or spatial shift of digit representation maps found | 5 |

## Materials and methods

### Participants

A total of 20 right-handed professional musicians took part in the study. All musicians fulfilled the following inclusion criteria: 1) had completed postgraduate musical training; 2) performed either as a soloist or ensemble player; 3) musicianship was their primary source of income. The patient group consisted of 11 musicians (10 male; mean age=49.9 years, SD=7.85). Patients were recruited via clinics at the National Hospital for Neurology and Neurosurgery and London Hand Therapy. Two neurologists (MJE and AS) with a special interest in Musician’s dystonia independently confirmed the diagnosis. For each patient, symptomatic fingers during musical performance with their primary instrument of choice were noted (guitar or piano). Symptomatic fingers were defined as (1) reported to be affected by patients and (2) had an objective deficit of motor control on examination by specialist (such as an abnormal posture, or recurrent pattern of abnormal movement on action). All patients had dystonic symptoms in the right-hand whist playing, two patients had bimanual symptoms (Table 2). The severity of overall impairment for each individual was quantified using the Tubiana and Chamagne scale.^20^ The control group consisted of nine healthy musicians with no history of musculoskeletal/functional impairment of the upper limbs (all male; mean age=41.0 years, SD=14.54). Both groups were matched for age (t_18_=1.75, p=0.097, unpaired t-test). The local ethics committee approved all study procedures and written consent was obtained from each participant according to the Declaration of Helsinki.

**Table 2.** Demographic and clinical details of musicians. *Age and Duration of symptoms are given indicated within ranges as per medRxiv guidance*. Mean duration of dystonic symptoms was 7.82 years (SD=7.21). All patients had symptoms in the right-hand (most commonly in the ring and middle fingers) and two also had dystonia of the left thumb. Severity was indicated by the score on the Tubiana-Chamagne scale and has the following possible values: 0-unable to play, 1-plays several notes but stops because of blockage or lack of facility; 2-plays short sequences without rapidity and with unsteady fingering; 3-plays easy pieces but is unable to perform more technically challenging pieces; 4-plays almost normally, difficult passages are avoided for fear of motor problems; 5-returns to concert performances). The average Tubiana-Chamagne score for the complete cohort was 2.55 (SD=0.688).

| Age | Gender | Instrument | Symptomatic fingers | Duration | Severity |
| --- | --- | --- | --- | --- | --- |
| 41-50 | M | piano | left thumb, right thumb | 1-10 | 2 |
| 41-50 | M | piano | left thumb, right middle, ring | 1-10 | 3 |
| 61-70 | M | piano | right ring, little | 21-30 | 2 |
| 51-60 | M | guitar | right index | 1-10 | 2 |
| 51-60 | M | guitar | right thumb, index | 1-10 | 2 |
| 41-50 | M | guitar | right middle | 1-10 | 3 |
| 51-60 | M | guitar | right middle, ring | 1-10 | 3 |
| 41-50 | M | guitar | right thumb | 11-20 | 2 |
| 51-60 | F | guitar | right index, middle, ring | 1-10 | 2 |

### Experimental Design

Patients and controls attended two independent study sessions (i) consent, explanation and practice (ii) performance of task in the MRI scanner. Two patients were unable to complete the task in the fMRI scanner due to anxiety/claustrophobia and imaging data acquisition could not be completed. In total data from nine musicians with dystonia and nine healthy musicians were analysed.

### fMRI experiment to measure finger-specific activity patterns

Data were acquired on a Siemens 3T TRIO MRI scanner with a 32-channel head coil. During finger stimulation, we measured the blood-oxygen-level dependent (BOLD) responses in both patients and controls. For each participant, evoked-BOLD responses were measured for passive and active movement conditions. To passively lift individual fingertips and record individual finger forces, we developed an fMRI compatible device with five piano-style keys. Each key had a small grooved circular platform onto which the fingertip could be placed. In the *passive* condition the circular platform was raised by the pneumatic piston so that the individual fingers of the right hand were passively extended by ∼15mm (Figure 1, for details, see Berlot et al. 2019^21^). We also added an *active* movement condition, during which participants used their left and right hands to performed individuated finger presses. In this paper we only report the passive movement condition, as it does not have any motor behavioural confounds and can therefore be strongly compared across patients and controls. Equivalent analysis of the active conditions, however, also did not show a difference between the two groups. The experiment began with participants fixating on a star in the centre of the screen. At the start of each trial, the fixation star turned a different colour for 1.36s to indicate one of three experimental conditions: 1) a white star indicated a *rest* condition, 2) a green star indicated the *active* condition, 3) a red star indicated the *passive* condition. A keyboard outline was presented to the participant with the instructed finger for the trial highlighted in green. During the *active/press* condition the star was replaced with a green letter ‘p’, which was the go-cue for participants to make a short isometric force press with the instructed finger. A force of 2.3N was required to register a successful key press following which the letter ‘p’ disappeared. The letter ‘p’ reappeared again after 1.5s to signal the start of the next press. This was repeated six times (total trial length approximately 10.68s). During the *passive* condition, the red star persisted and the indicated finger was lifted 6 times for 1 sec with pauses making a common total trial length of 10.5s. Participants were instructed to relax fingers in both hands during the *rest* condition. Instructed fingers for each trial were selected in a pseudo-random order, such that all possible combinations were tested twice in each run (10 active press conditions with 2 hand and 5 fingers, and 5 passive movement conditions for the fingers of the right hand only). In addition, 3-5 rest conditions of varying lengths (14.9s, 25.5s or 36.1s) were randomly interspersed within each run. During each functional run, 126 images were obtained at an in-plane resolution of 2.3mm × 2.3mm (2D echo-planar sequence, TR=2.72s, 32 interleave slices with thickness=2.15mm and gap=0.15mm, matrix size=96×96). The first three images in the sequence were discarded to allow magnetisation to reach equilibrium. Field maps were obtained and used to correct for field strength inhomogeneities ^22^. Finally, a T1-weighted anatomical scan was obtained (3D gradient echo sequence, 1 mm isotropic, field of view=240 × 256 × 176mm).

**Figure 1.**
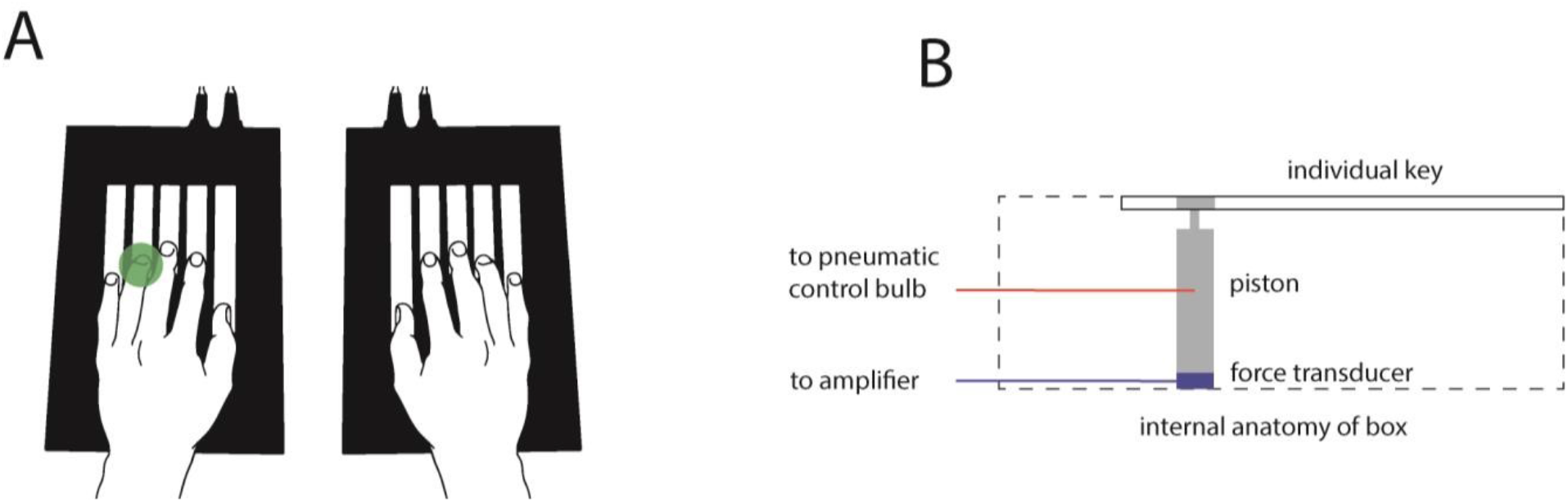
Equipment for Experiment. A. Our custom-built keyboard had force transducers and pneumatic pistons embedded within each key. B. Schematic diagram of the internal anatomy of keyboard.

### Imaging analysis

The functional imaging data from each participant was minimally pre-processed using SPM tools.^23^ The data were corrected for slice timing, realigned to correct for head motion across runs, and co-registered to each participants’ T1-image. The pre-processed functional data was then analysed using a generalised linear model, with a separate regressor for each trial condition/finger/run. The evoked-BOLD activations for each condition were modelled using a boxcar function (duration=8.7s) and convolved with a standard hemodynamic response function. The parameter estimates for each run and condition was divided by the root mean-square error from the first level model to obtain a t-value for the condition>rest contrast. These t-values were then used to investigate the organisation of finger maps in primary sensory and motor cortex. Each participant’s T1-image was used to reconstruct the pial and white-grey matter surfaces using Freesurfer.^24^ The surfaces are meshes that consist of nodes that are connected with edges. Individual surfaces were inflated to a sphere and then registered across participants by matching them to a common template (fsaverage) using the sulcal-depth map and local curvature as minimisation criteria. Two regions of interest (ROIs) were defined on the group surface using probabilistic cyto-architectonic maps aligned to the average surface.^25^ Surface nodes with the highest probability for Brodmann area (BA) 4 1.5cm above and below the hand-knob were selected as belonging to the hand area of primary motor cortex (M1). Similarly, nodes in the hand-region in BA 3a and 3b, again 1.5cm above and below the hand knob were selected for the hand area of primary somatosensory cortex (S1). We did not consider BA 1 and 2, as finger representations tend to be more overlapping and less clearly organised even in healthy controls.^26^ To avoid possible contamination of signals across the central sulcus, we excluded all voxels that had more than 25% of its volume located on the opposite side of the sulcus. All other voxel that were partly positioned in the between the pial and white-grey matter surface in the two ROIs were used in the analysis.

### Quantifying finger representations using spatial analysis

Previous studies have reported that the spatial distances between different finger representations were significantly reduced in dystonia. Motivated by these studies, we employed different spatial and multivariate measures to characterise the activity pattern in S1 (BA3a and BA3b) and M1 (BA4) to determine whether patients showed an altered structure of finger representations relative to controls. We analysed the activity maps for passive finger movements on the right hand (symptomatic in musician’s dystonia). The t-values for each finger were projected onto a flattened version of each individuals’ surface. To analyse the spatial layout of the finger representation we employed a number of different methods to seek replication of previous studies. For the first method, we simply used the location of the maximal activity for each finger (see Figure 3) within the predefined ROI (see above). In the second approach, in order to take into account the entire activated region, we used the centre of gravity (COG), the average x and y location of each surface vertex, weighted with *w*_i_.

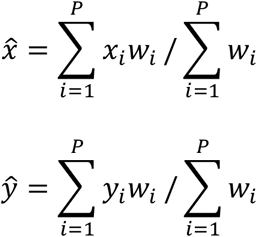

In the case for the linearly-weighted COG, we set *w*_i_ to the t-value for positive activations, and to 0 for negative activations. Finally, to obtain different compromises between the maximal activation and the COG approach, we also calculated the COG, using a weighted softmax function ^27^ of the t-value (*t*_*i*_) as *w*_*i*_.

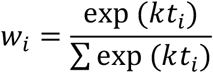

For a softmax parameter *k=0*, all surface nodes would have the same weight and the resultant coordinate would be the COG of the entire ROI. For larger value of k, the weight of the more highly activated surface nodes will be much higher than for the other nodes. In the extreme of *k* → ∞, the coordinate will simply reflect the location of the maximal activation. Thus, by varying *k* over multiple values (0.05, 0.1, 0.2, 0.4, 0.8, 1.6, 3.2) we were able to explore analyses that either took into account the entire activated area, or concentrated on the areas of highest activation (see Figure 4). To assess the ability of each metric to reveal the somatotopic ordering of the finger representation, we performed a MANOVA for each group, using the x- and y-coordinates for each finger as dependent measures. The MANOVA test for any systematic differences between the spatial locations of any pair of fingers. To test group differences in the spatial layout of digit representations, we calculated the Euclidian distance between the 10 possible pairs of digits. We then compared the average distance between groups using a Student *t*-test for independent samples. We also assessed for differences in the relative spatial layout by submitting the 10 distances to a repeated measures ANOVA and assessing the group x digit pair interaction using an *F*-test.

### Quantifying finger representations using representational similarity analysis

As an alternative to spatial measures of finger representations, we also employed representational similarity analysis (RSA), which assesses the similarity between different activity patterns in the ROI, while ignoring the spatial arrangement.^28, 29^ The beta-weights from the generalised linear model were extracted to obtain the finger-specific activity patterns in the ROIs. The dissimilarity between the activation patterns for each finger pair (z_i_, z_j_) was computed using the cross-validated Mahalanobis distance.^30^ The voxel-by-voxel covariance matrix Σ was estimated from the residual from the first-level model and regularised by shrinking all off-diagonal elements towards zero.^31^ An unbiased estimate for the squared Mahalanobis distance can then be calculated as:

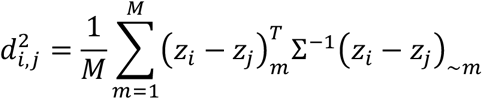

For each of the M runs, the patterns are estimated from the data from that run (m) or from all other runs (∼m). This cross-validation procedure ensures that the expected value of d is zero if two patterns are not different from each other and that the distance estimates are unbiased.^32^ We computed distances between all 10 pairwise combinations of fingers, separately for each active/passive movement condition, for each hand, and within each ROI (S1 and M1). Statistical group differences in the average distance and relative arrangement were tested as described for the spatial distances. The relative arrangement of the digits can also be visualised in a two-dimensional representational space using classical multi-dimensional scaling.^33^

### Surface-based searchlight analysis

To detect possible overall differences in the location or spatial extent of the digit representation, we also conducted a searchlight analysis.^34^ Based on the individual cortical surface reconstruction, we selected - for each surface node - a circle on the surface that contained 60 voxels between the pial and the white-matter surface. For each of these searchlights, we computed the average cross-validated Mahalanobis distance across all 10 pairs of fingers (see above). The average dissimilarity was then mapped back to the node in the centre of the searchlight. By repeating this process for each surface node, we build up a cortical map of cortical regions that contained digit information for each participant. The maps were then compared using a *t*-test for independent samples and uncorrected threshold of *t*_15_= 3.2860, *p*<0.005, and corrected for cluster size using Gaussian Field map correction.^35^

## Data availability

Data and analysis code are available on request from the authors.

## Results

We wanted to determine whether there were measurable alterations of the finger representations in the sensorimotor cortices in Musician’s dystonia. We therefore used high-resolution fMRI to measure evoked-BOLD responses in S1 and M1 during passive stimulation and active finger presses in our cohort of participants (see methods).

As expected the activity patterns for each finger press were distributed with substantial overlap between the different fingers in both pre- and post-central gyrus, even in healthy musicians (Figure 2). ^17^ Importantly, there was substantial variability in finger-specific activity patterns across individuals. Motivated by previous papers^6, 8, 9, 11, 14, 36^, we first considered spatial measures to characterise the digit representations in S1 and M1. Given this somewhat fractured nature of the digit activation patterns, it is not immediately clear how to summarise the spatial relationship between the finger representations. We therefore employed a range of methods to determine the location of the representation of each finger. We used either the location of maximal activation (Figure 3a, star) or the centre of gravity of the activated vertices, weighted by the size of the activation (Figure 3a, cross). To explore a wider range of intermediate methods, we applied a softmax approach (see method). By varying the softmax parameter k, we can weight each positively activated vertices equally (*k*=0), or only use the most highly activated vertex (*k* → ∞). As can be seen in Figure 3a, the variation allows a trade-off between taking into account the entire activated regions and concentrating only on the most active regions. The Euclidean distances between the five COG estimates for the different fingers gave 10 pairwise distances, which in turn quantified the spatial geometry of finger representations in the hand-knob for each individual.

**Figure 2.**
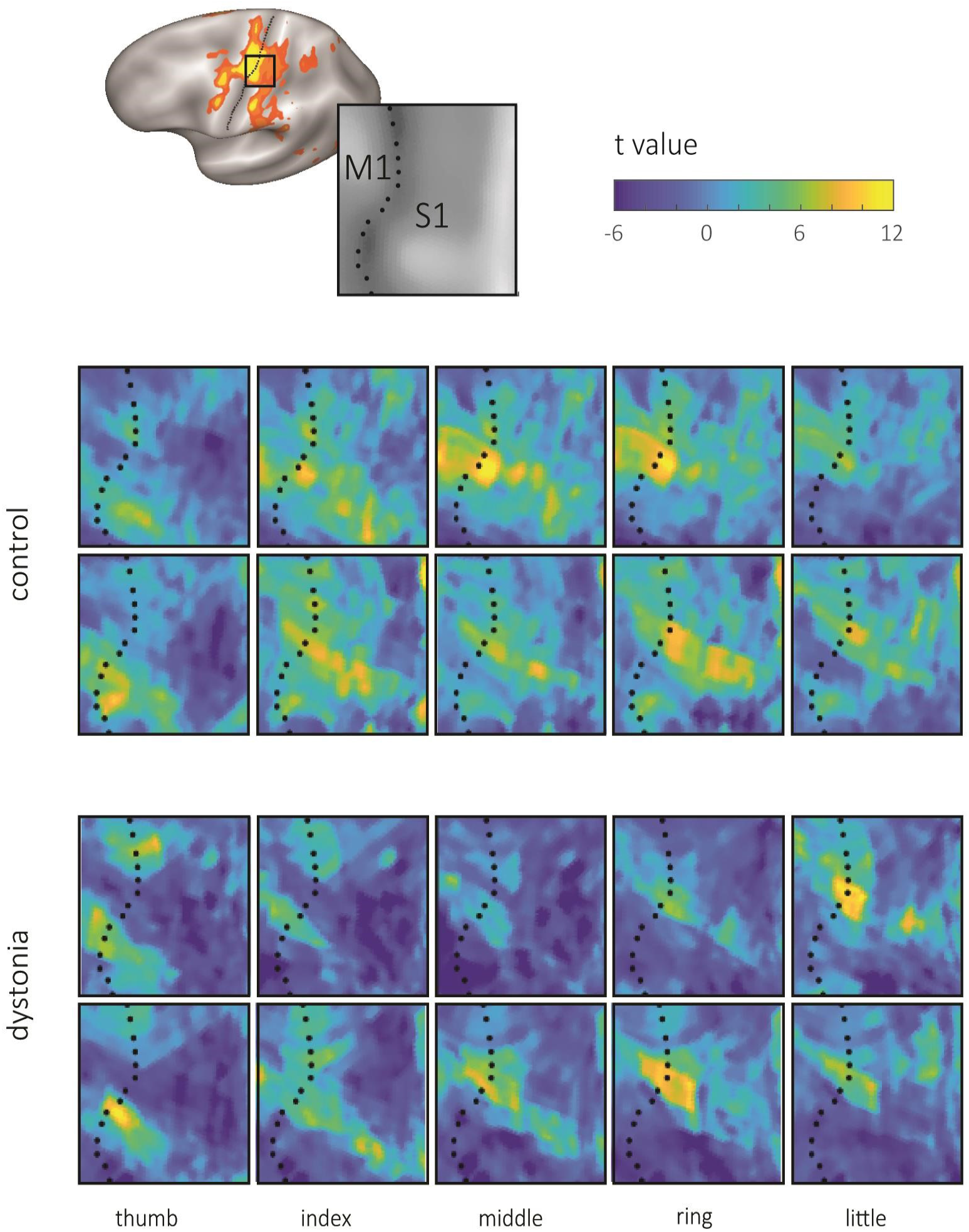
Individual activity patterns of finger representations in the left sensorimotor cortex during passive extension movements of fingers of the right hand. Each row shows the activity patterns from a single individual; two healthy musicians and two musicians with dystonia. Note the considerable variability of finger-specific activation patterns across participants.

**Figure 3.**
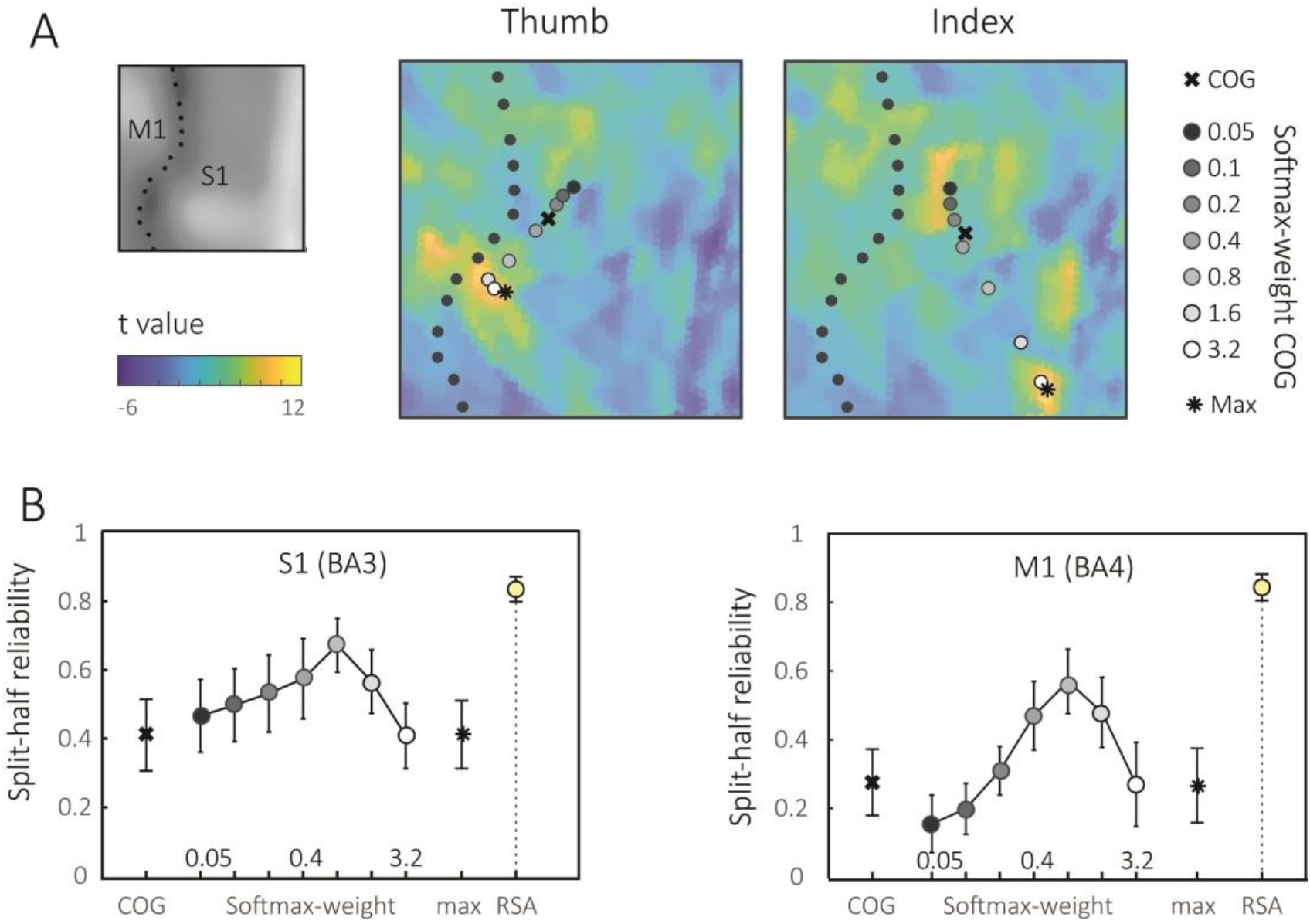
Comparison of different methods to characterise the spatial layout of digit representations. **(A)** Activity pattern (t-values) for the thumb and index finger of a selected control participant over the region of interest S1. Star: the location of peak activation on each map (max). Cross: the centre of gravity (COG) of the activated regions, weighted by the t-values for each vertex. Circles: softmax-weighted COG, with a k-parameter of 0.05 (close to COG of the region) to 3.2 (close to max). **(B)** Split-half reliability of the distances between digit representations for COG, softmax-approach, peak activation (max), and representational similarity analysis (RSA) for S1 and M1.

**Figure 4.**
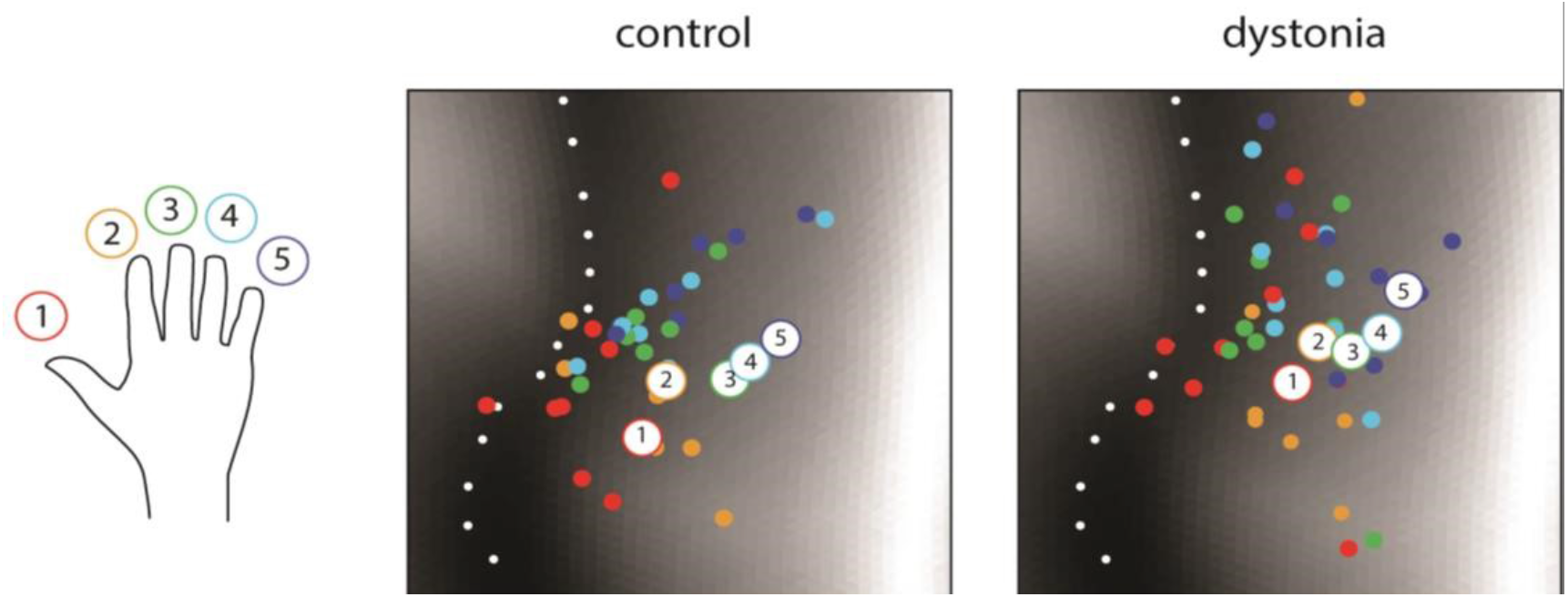
Somatotopic order of finger representations in S1. Using the optimised softmax-weighted COG (k-parameter = 0.8) a somatotopic arrangement of fingers was demonstrated at the group level. Large white circles indicate group mean, and small colored circle the individual variability: thumb = red, index = orange, middle = green, ring = pale blue, little= dark blue. Locations are shown on the same flattened cut out of the cortical surface as used in Figure 2 and 3. The location of the central sulcus marked by a dotted line.

To decide between these different methods of characterising the spatial layout, we determined the split-half reliability of the 10 pairwise distances between digit centres, using odd and even runs from each participant. Both the COG method, as well as the point of maximum activation led only to a mediocre within-subject reliability, with split-half correlations around 0.4 for S1 and 0.3 for M1. The softmax approach performed best, with a value of k=0.8 providing a best compromise between the entire region of activation and the most highly activated regions. For all subsequent spatial analyses, we focused therefore on this metric.

For both controls and patients, the corresponding softmax COG estimates uncovered a clear somatotopic order of finger representations in S1 (MANOVA, controls: *χ*^2^_8_=34.82, p=2.879e-05; patients: *χ*^2^_8_= 23.28, p= 0.0037). Using this metric (and any other spatial measure computed), however, we found no differences in the average distances between digit centres (S1: *t*_15_=0.070, *p*=0.945, M1: t_15_=0.427, *p*=0.676), nor in the relative spatial layout (S1: *F*_9,135_=1.0864, *p*=0.3769; M1: *F*_15,135_=0.8003, *p*= 0.6166). Taken altogether, the extent and exact geometry of somatotopic ordering of finger representations was indistinguishable between healthy musicians and musicians with dystonia.

Spatial measures to quantify finger representations have significant weaknesses. Consider for example the activity patter for the index finger shown in the first row of Figure 2 which shows four different small cluster of activity in S1. Any spatial measure summarises these clusters into one location, therefore providing only a poor description of the complexity of the underlying map.

To overcome this limitation of spatial metrics, in our second analysis we used multivariate pattern analysis to quantify finger representations. For both patients and controls, we estimated the cross-validated Mahalanobis distances (see methods) between all pairs of evoked-activity patterns during passive stimulation of the fingers in the right hand (Figure 5). Dissimilarity structures for controls and patients could be visualised as vector plots and along two dimensions using multidimensional scaling (Figure 5a, see methods). Both plots illustrate the typical structure of finger representations in the sensorimotor cortex with largest dissimilarities for thumb and ring fingers and smallest for the middle and ring fingers (Ejaz et al., 2015). The split-half reliabilities for RSA measures were above 0.8, significantly higher in both S1 and M1 than any of the spatial measures (Figure 3b).

**Figure 5.**
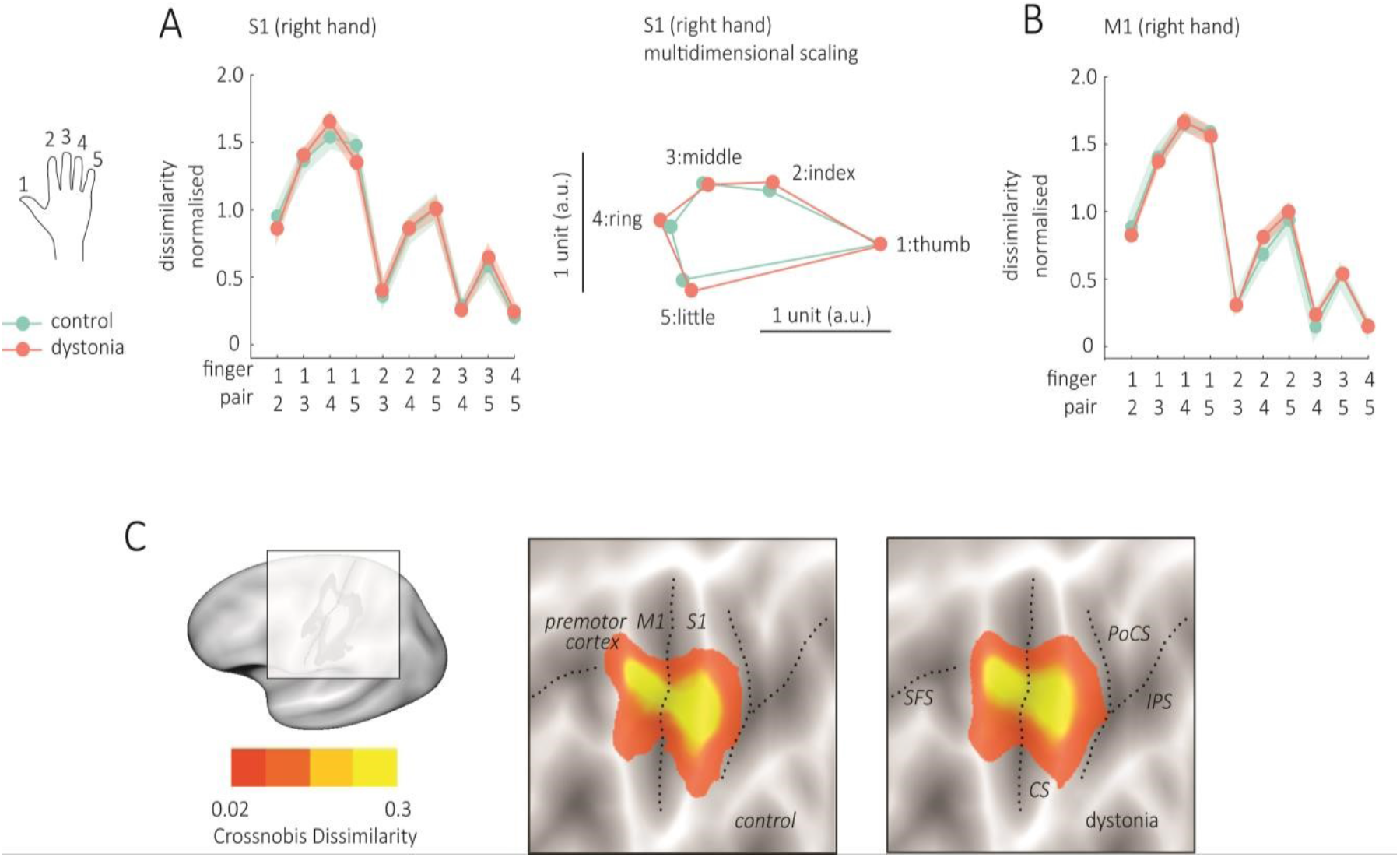
Representational structures for passive, extension movements are not altered in Musician’s dystonia. Pairwise cross-validated Mahalanobis distances between activity patterns for passive finger movements were estimated to get dissimilarity structures in S1(A) and M1(B). Values close to zero indicate the patterns are similar, while unique patterns are indicated by higher values. Dissimilarity structures were highly similar for patients and controls, in S1 and M1 and the uniqueness of the thumb activity pattern from other patterns is easily visualised by projecting the dissimilarity structures into a low-dimensional space using multidimensional scaling. C. The extent and location of digit representations for healthy musicians (controls) and musicians with dystonia was also explored. The average dissimilarity between digit-specific activity patterns is shown on an inflated version of the lateral left hemisphere in two rectangular panels. The location of the superior frontal sulcus (SFS), central sulcus (CS), postcentral sulcus (PoCS) and intra-parietal sulcus (IPS) are indicated by dotted lines. Gray scale in the background indicates cortical folding (dark = sulcus, light = gyrus). The approximate mapping of the rectangular panel to the left hemisphere is illustrated and the average dissimilarity value indicated by colour bar: low dissimilarity red, high dissimilarity yellow.

However, even when using the RSA dissimilarities between finger-specific activity patterns, we found no significant difference between the groups – neither in the average dissimilarity (S1: t_15_=0.266, p=0.794, M1: t15=-0.089, p=0.931), nor in the pattern of dissimilarities (S1: *F*_9,135_=0.26, *p*=0.9831, M1: *F*_9,135_=0.03, *p*=0.999).

Finally, we asked whether we could detect any expansion or spatial shift of the entire digit representation across the sensory-motor cortex. Figure 5C shows a surface-based searchlight map of places on the lateral hemisphere, were the local activity patterns differed between different fingers. For both healthy control musicians and musicians with dystonia, the hand area of M1 and S1 are clearly visible. A statistical comparison between the two maps (see methods), however, did not yield any significant clusters of increase or decrease distances in the patient group.

To summarise, even when using an optimised measure of pattern organisation, we were unable to detect any alteration of basic finger representations in primary sensorimotor cortex between musicians with and without dystonia. Neither the overall location nor extent of the digit representation (Figure 5C), nor the arrangement of the digit-specific patterns within this region (Figure 5A-B), showed any group difference. Overall, our results contradict previous reports that argue that abnormal finger representations in primary sensorimotor cortex are the cause for the loss of finger control in musicians with dystonia.

## Discussion

In this study, despite careful technical work, our fMRI data did not provide any evidence for an alteration or distortion of finger representations in primary somatosensory or motor cortex. Our results challenge the hypothesis that the task-specific dystonias are caused by an alternation of the somatotopic organisation of representations of the hand. Instead, we offer the alternative hypothesis that task-specific dystonia is encoded within a hierarchical skill network which has implications for how we design and prioritise future rehabilitation strategies.

The original evidence for somatotopic disruption in task-specific dystonia came from work in primates required to perform repetitive hand movements (Figure 6A).^37^ One limitation of this experimental paradigm as a model for Musician’s dystonia is that monkeys were trained on repetitive whole hand grasping movements which are likely controlled differently from the fractionated movements of individual fingers akin to musical performance.^38^ Furthermore, from a conceptual point of view, it has always been difficult to define how abnormal S1 maps translate into a task-specific motor deficit. Conceptually, if the sensory representation of fingers were less differentiated in S1 for any finger, the corresponding blurring of incoming sensory data would presumably affect all manual tasks. Overall, therefore, it seems unlikely that causal neural engrams for dystonia are “hardwired” into generic hand maps within S1 or M1.^39^

**Figure 6.**
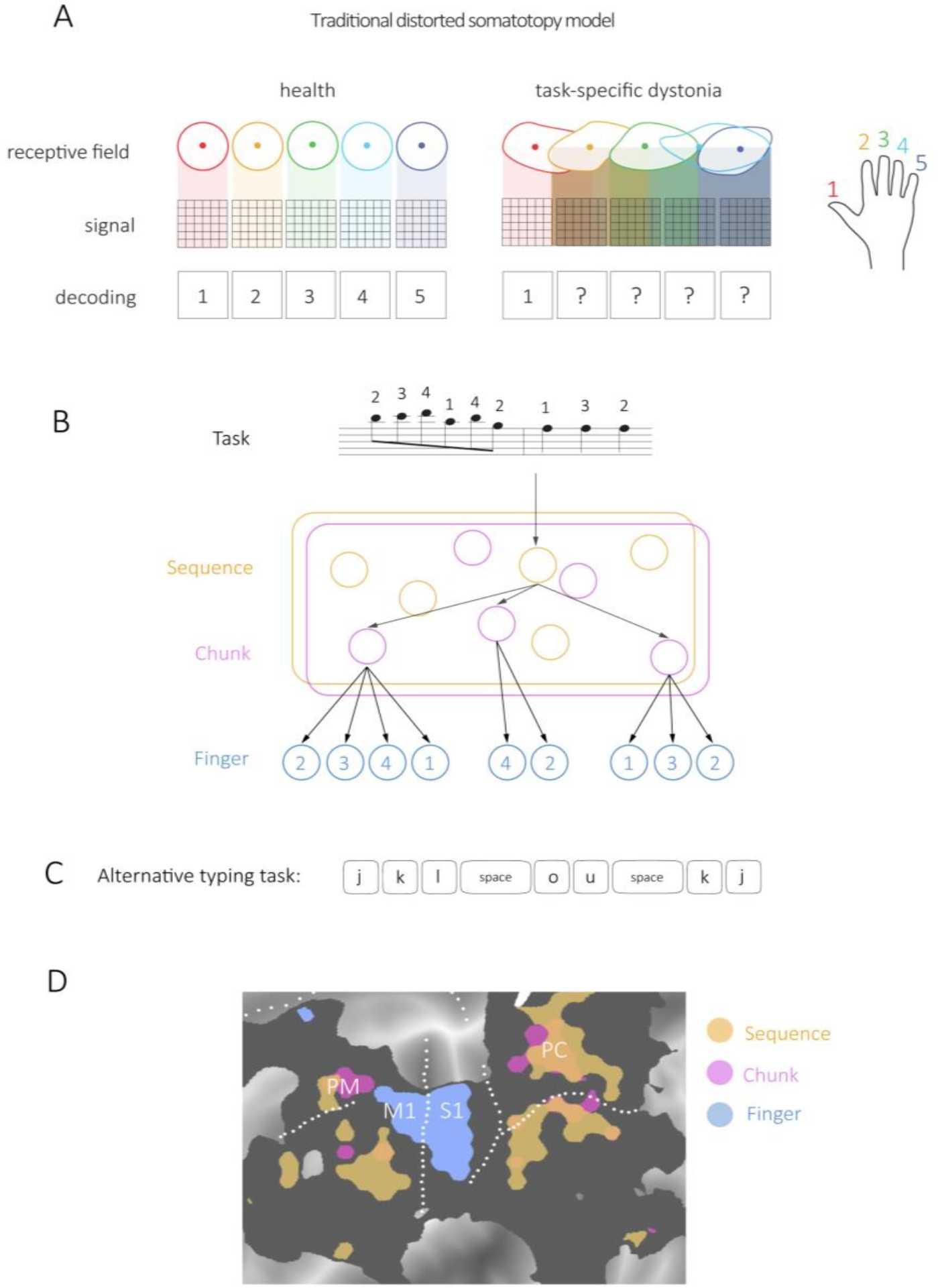
Disease models for task-specific dystonia. A Traditional distorted somatotopy hypothesis. The receptive field for each finger in S1 is drawn (thumb=red, index=orange, middle =green, ring=light blue, little=dark blue). In health, this idealised cartoon shows receptive fields as discrete areas. The corresponding signal across a strip of cortex cleanly maps to an individual finger. In task-specific dystonia overlapping and disorganised receptive fields lead to a mixing of signals and uncertainty of finger mapping. B. Possible neural representation of a complex task. A small piece of music, which requires a series of finger presses of the right hand on a piano, may be represented in a hierarchical fashion with sequences and chunks (grouped 4-2-3 in this example) that then in turn activate the elementary movement components. C. An alternative task such as a series of keyboard presses requires the same sequence of finger presses as the piano task. However, if the chunking converges on a different pattern (e.g. grouping of 3-3-3), immediately a different set of higher order control elements are required for typing the same sequence of finger presses. This is one potential neural mechanism for a taskspecific deficit. D. Recent fMRI work in normal controls shows that while single fingers are represented in M1 and S1 (blue), hierarchically more complex chunks (pink) and sequences (orange) are represented in overlapping regions of premotor and parietal cortex^41^.

Methodologically, there are also limitations with experimental arguments used to support the somatotopic disruption model. In human studies, the overlapping nature and complexity of the activation patterns makes it difficult to find good metrics that summarise their organisation. We have therefore systematically explored a range of methods that have been used in the literature. We found (in this work and previous studies) that spatial centre-of-gravity and location-of-maximal-activation metrics have low reliability within and across different healthy musicians, and hence failed to reveal an invariant organisation of finger maps that would be characteristic for a neurologically healthy individual. Given their failure to do this, it is over-optimistic to think that they should reveal systematic deviations from the healthy pattern in disorders such as task-specific dystonia. Representational similarity analysis, in contrast, was confirmed to uncover representations that were highly robust within and between individuals.^17^ Our passive finger lift condition focused on proprioceptive input related to finger movement and the active finger press corresponded to muscle activation patterns elicited during near isometric finger presses. However, despite the different behavioural features, both tasks elicit highly similar activity patterns in S1 and M1^40^ and neither of these task conditions revealed changes in representational architecture.

These results therefore suggest the neural architecture of basic digit representation of within S1 and M1 is intact, and that the disruption of representations must occur in other, possible higher-order areas. Such a stance would fit with the neuroscientific literature that examines skill encoding in which the motor hierarchy recruits higher order regions to represent task-specific features such as the order in which fingers are required to move within a sequence. For example, in a study in which volunteers were trained to learn a sequence of finger presses intensively over five weeks with four interval scans; participants became faster and more accurate at executing the sequences and this learning was associated with the stabilisation of sequence specific-activation patterns in premotor and parietal areas.^21^ Furthermore, when probing the representation in different regions, it becomes clear that individual movements are represented in primary motor and sensory cortex (Figure 6B), while encoding of movement *sequences* as either chunks (short stereotyped elements) or longer sequences were only found in premotor and parietal cortices.^41^ Given that individual finger presses are mostly intact in tasks removed from musical performance, the features of higher order task-specific networks may therefore be better placed to explain the phenomenology of task-specificity dystonia (Figure 6B-D).

In the future, therefore, we require more sophisticated analytical and experimental methods to gain further insight into the aetiology and disruption of representations in task-specific dystonia. In terms of aetiology, the recurrence of certain risk factors across different individuals in task-specific dystonia does suggest that there may be shared neural vulnerabilities at a group level. For example, over-trained skills are likely to have features of encoding that are distinct to every day skills such as extended planning horizons/sequences and a narrow capacity to generalise to other tasks of shifts in the original tasks demands.^42, 43^ What is clear is that our results motivate the search for the neural correlates of task-specific dystonia away from generic digit maps in primary sensorimotor cortex. Such a view also fits with an expanding literature which emphases that task-specific dystonia is associated with changes across a broad network and that compared to other subtypes of dystonia there is greater involvement of sensorimotor cortical areas such as the premotor and parietal cortex^44^.

Our results have repercussions for how we treat task-specific dystonia. Early retraining methods that developed in line with the distorted somatotopy hypothesis were optimistic that sensory super-training (such as learning braille) could be helpful with the idea that improving sensory discrimination of the affected hand would lead to remapping and normalisation of dystonic digit maps within somatosensory cortex.^45-47^ However, such retraining therapies are not widely used in clinical practice and are not reliably effective^48^. Our findings offer an optimistic therapeutic starting point as we believe that the neural representation of incoming sensory information related to passive lifts of individual fingers and the representation of the execution of individual finger presses is intact. Our results therefore encourage therapeutic interventions that specifically tap into task-specific sensorimotor interventions (e.g. sensorimotor retraining and differential learning).^49^

Of course, a major limitation of our study is that our main conclusions rest on a null-result, which could also have been caused by insufficient sample size or insensitivity of fMRI to true alterations in the organisation of basic finger representations. A Bayesian analysis or equivalence testing is made very difficult, as we have no proper basis to estimate the size of the likely effect that we should have observed. In favour of our conclusion, however, we have carefully evaluated the different methodologies previously used to show alterations and found that these techniques are generally rather unreliable. In contrast, the most robust technique available to assess neuronal population activity patterns provide substantial evidence that the basic structure of basic finger representations is normal in these individuals.^29, 32, 50, 51^ Alternatively, we may have not captured neural differences due to the task tested. It is likely that dystonia needs to be clinically manifest in order for the abnormal skill network to be activated. However, it is worth emphasising that none of the studies that have reported altered somatic finger representations in the past have used a task that generated dystonia.^7-14^ We therefore believe the identified shortfalls with the distorted somatotopy model are significant and valid.

In summary, in this study we found no evidence of abnormal finger representations in primary sensorimotor cortex using fMRI and multivariate pattern analysis. Our data support the development of alternative disease models that have the advancement of novel therapeutic interventions at their core.

## Data Availability

Data produced in the present study are available upon reasonable request to the authors

## Acknowledgements

We thank the musicians with and without dystonia that generously gave their time for these experiments.

## Supplementary material

**Supplementary Figure 1.**
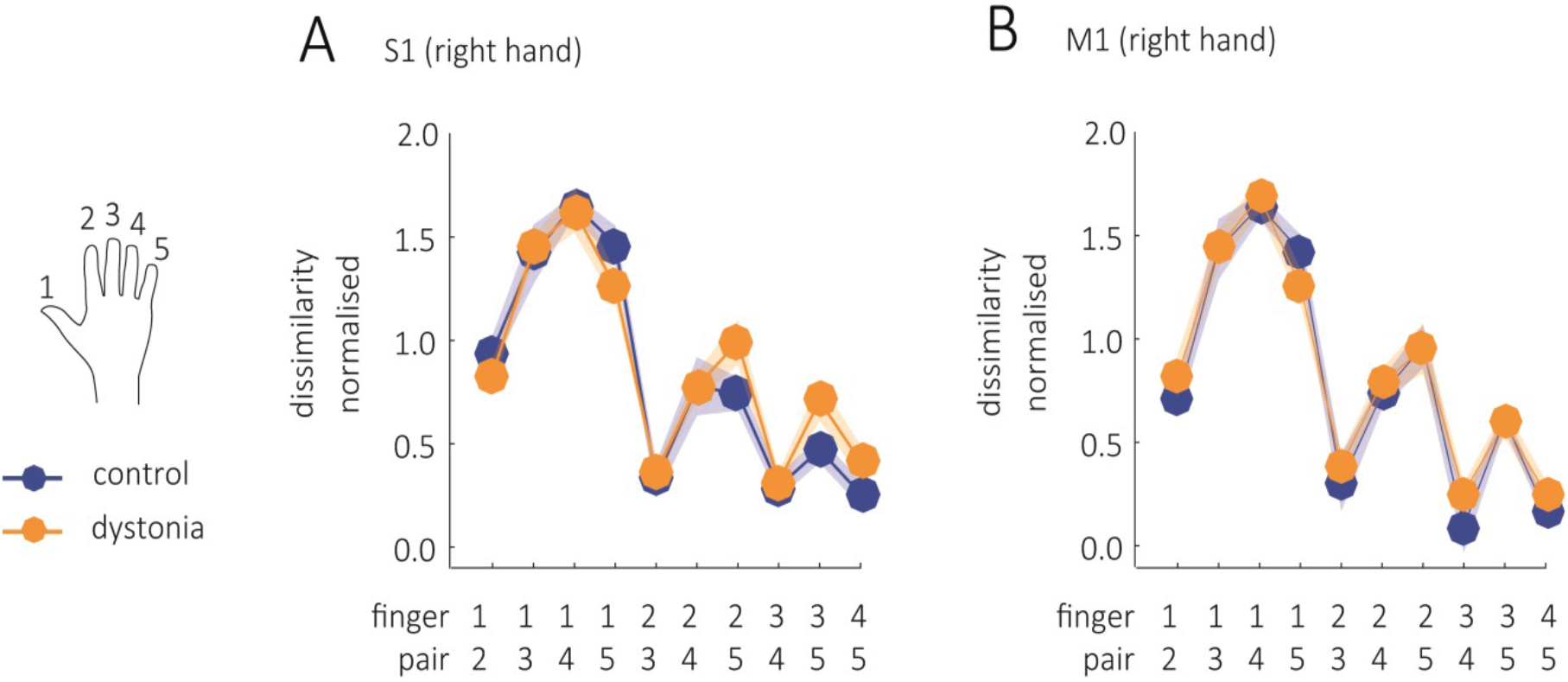
Representational structures for active finger presses are not altered in Musician’s dystonia. Pairwise cross-validated Mahalanobis distances between activity patterns for finger presses were estimated to get dissimilarity structures in S1(A) and M1(B). Values close to zero indicate the patterns are similar, while unique patterns are indicated by higher values. Dissimilarity structures were highly similar for patients and controls, in S1 and M1.

